# Women’s autonomy in consent and debriefing for caesarean section: a thematic analysis of healthcare providers’ views and women’s lived experiences and expectations in 20 hospitals across West Cameroon

**DOI:** 10.1101/2025.10.29.25339125

**Authors:** Jovanny Tsuala Fouogue, Miho Sato, Louise Tina Day, Mitsuaki Matsui, William Carter Djuatio Kenne, Bruno Kenfack, Lenka Beňová, Veronique Filippi

**Author notes:** **Corresponding author** Jovanny Tsuala Fouogue, (JTF).

## Abstract

Women’s autonomy in providing informed consent for caesarean section (CS) and receiving post-operative debriefing is essential for a high-quality, women-centered experience of care. Yet, in many low-income settings, while the clinical quality of CS is known to be suboptimal, evidence on its woman-centredness remains limited. Our study examined women’s autonomy around CS in 20 hospitals across nine health districts in the West Region of Cameroon. Between March 2024 and August 2024, we conducted in-depth interviews with 69 CS healthcare providers and 20 women within 30 days of a CS as well as 20 focus group discussions with 128 pregnant women attending antenatal clinics. Data were analyzed using inductive coding and thematic analysis of verbatim transcripts. Five themes reflected barriers to women’s autonomous decision-making in consent for CS and post-operative debriefing interactions. These were: embeddedness in a patriarchal and collectivist society; primacy of the emergency situation in CS decisions; medical paternalism and women’s deferential behavior; financial and logistical model of CS care; and weak regulatory framework for clinical practice. The sixth theme delineated a bold emancipatory counter-current, challenging the afore-mentionned barriers and promoting women’s autonomy. Our results suggest that policymakers should promote consent guidelines that support women’s agency and ensure debriefing focusses on their needs and social support. Community engagement should also address the deeply rooted influence of marital norms and women’s limited financial resources in CS decision making.

## Introduction

Autonomy is defined as self-determination or self-governance and conveys the notion of unrestricted ownership of oneself in thought, in will and in action (Nemie et al., 2014; Stoljar and Voigt, 2022). Alongside “beneficence”, “non-maleficence” and “justice”, autonomy is one of the four moral principles in health care since their popularization by Beauchamp and Childress in 1979 - a decade after the Belmont Report established them as core protections for human subjects in biomedical and behavioral research (Beauchamp and Childress, 2009; US Department HHS, 1979). Of those four pillars autonomy has changed the most overtime, reshaping physician-patient relationships. Now widely endorsed by medical and regulatory bodies across cultures and countries, patient autonomy was once explicitly rejected - as illustrated by this extract of the 1847 American Medical Association code of medical ethics: “*The obedience of a patient to the prescriptions of his physician should be prompt and implicit. He should never permit his own crude opinions as to their fitness, to influence his attention to them*.” (5,6). Informed consent or refusal is the instrument through which patients exercise autonomous decision regarding physician-recommended treatments (7). To be valid and effective, informed consent must encapsulate patient’s competence and voluntariness, based on the provision and understanding of relevant information, and a notification of decision (5).

Bodily autonomy and reproductive rights have been central to women’s emancipation movements which, through confrontation with patriarchy norms and power structures, have shaped diverse socio-political and legal gender landscapes across the globe (UN, 2019). Those rights remain a cornerstone of the global gender equality agenda, enshrined in Sustainable Development Goal 5: “*Achieve gender equality and empower all women and girls”*. Yet, progress remains uneven and slow. At the current pace, SDG 5 will not be achieved by 2030 with significant disparities between and within regions and countries (9). This includes gaps in areas such as Target 5.6.1 which tracks the “proportion of women aged 15 – 49 years who make their own informed decisions regarding sexual relations, family planning and reproductive health care”.

Clinical services for reproductive health need to ensure that health facilities are conducive settings for women’s rights. The World Health Organization (WHO) identified restrictions on women’s autonomous decision-making - such as requiring consent from parents, spouses or guardians for treatment including the absence of informed consent - as discrimination against women in access to care (WHO, 2017). Any restriction on access and quality of reproductive health, including maternal health services, raises the burden of ill-health and must be documented and adressed (11,12).

Most maternal and perinatal deaths and severe complications occur because women cannot access high quality care around the time of birth (13–15). Cesarean section (CS) has substantial benefits for the health of women and their babies, when medically indicated. Although CS is the most commonly performed surgical procedure for women worldwide, access to it and its quality remain limited in many low-and middle-income countries (LMICs) (16,17).

Current quality of care frameworks recognise experience of care as essential for achieving women-centred outcomes (US IOM, 2001; Tunçalp et al., 2015; US AHRQ, 2025). The three dimensions of experience of care - emotional support, respect and dignity and effective communication - are part of the WHO quality of care framework, and align with the core elements of relational autonomy in moral philosophy (1). While moral philosophy focuses on the socio-political dynamics of autonomy, the bioethical and medical spheres mainly engage with autonomy through informed consent (IC) (7). IC before CS and debriefing (post-procedure counselling, which provide complementary information) are critical to improve the acceptability of the procedure in LMICs where it remains dreaded and rejected (21)

In Cameroon, more than half of women in couples reported not being involved in household decisions regarding their health care and high gender inequality, patriarchal norms and power structures greatly restrict women’s autonomy (22,23). Unlike for abortion and contraception women’s autonomy for CS consent and debriefing has received little attention in the country’s guidelines for reproductive health (Ministry of Public Health - Cameroon, 2018).

In Cameroon, health system constraints include persistent severe shortage in human resource for health. In 2022, the physicians-to-population and nurses-midwives-to-population ratios were 1.36 and 6.59 per 10,000 population respectively (25,26). Like most services, CS is mainly funded by out-of-pocket payments with only 6% of the population having a health insurance in April 2023 (27,28). Women die in large numbers during childbirth. The maternal mortality ratio was 258 per 100,000 live births in 2023 (29).

Given the unique context of African settings – with high maternal mortality, limited access to life-saving CS, and substantial barriers to reproductive health decision making – our study explored the dynamics of women’s autonomy during consent for CS, and debriefing interactions in West Cameroon.

## Material and Methods

### Ethics statement

This study was ethically approved by the research governance and integrity office of the London School of Hygiene & Tropical Medicine (Reference: LSHTM Ethics Ref: 29898) and the Regional Ethics Committee for Human Health Research in the West Region of Cameroon (Reference: N° /984/25/10/2023/CE/ CRERSH-OU/VP). All participants provided written informed consent prior to data collection and minors were not included. Data were anonymised and handled confidentially.

### Study design

#### Qualitative approach, guiding theory and research paradigm

We conducted a cross-sectional qualitative study of CS providers’ views and women’s lived experiences and expectations. We captured data first through IDIs to explore women’s experiences of IC and debriefing because we dived deep into personal health histories and privates lives. We also conducted IDIs with CS providers (nurse-assistants, nurses, midwives, surgeons, obstetricians, scrub nurses, anesthetists), a diverse group of specialists for whom IDIs were well-suited to explore the effects of complex multidisciplinary and hierarchical structures as they can minimise the influence of professional power dynamics on openness. Second, we used FGDs to garther pregnant women’s expectations after antenatal clinic, to foster interactions and refinement of ideas, without going into personal sensitive matters.

We chosed interpretivism as the research paradigm as it stresses individual agency and interpretation within social structures and institutions. Interpretivism upholds that truth and knowledge are subjective as well as historically and cuturally situated, based on people’s experiences and their understanding of them (30). This research paradigm suits the pioneering nature of our research, in West Cameroon and its exploration of the negotiated nature of autonomy in CS consent and debriefing.

### Regional context and study sites

#### Regional context

The West Region is one of ten administrative regions in Cameroon serving a population of 2’398’224 inhabitants. (31,32) (33). In 2018, the total fertility rate was 5.5 and 97% of childbirths were facility-based while in 2022-24 the CS rate stood at 10% with 80% being emergency procedures (34) (35). Direct out-of-pocket payments for CS remain prohibitive and caused catastrophic health expenditures in 42% of cases in two neighboring regions in 2023 (27). Despite a high regional literacy rate, gender inequality index is high and patriachy is deeply rooted in the collectivist social fabric of communities (29,36). The region is home to semi-bantou ethnic groups, the largest being the Bamileke followed the Bamoun.

#### Study sites

Across the 20 health districts, CS is conducted in 96 hospitals. We included all twenty hospitals that performed ≥100 CSs in 2022, spanning nine health districts.

#### Participant selection

We recruited participants from March 4, 2024 to August 29, 2024. We purposively selected : pregnant women attending antenatal clinics, postnatal women discharged within thirty days of a CS (hereafter “post-CS women”), their accompanying relatives on the day of the CS when relevant, and CS providers. We invited pregnant women face-to-face to participate in in-person FGDs. We invited post-CS women by phone and conducted face-to-face IDIs at a location of their choice outside hospitals. During the calls, we asked post-CS women who reported limited involvement in consent interactions to indicate their accompanying relatives whom we also included with their agreement. We conducted face-to-face IDIs with CS providers at their workplaces. We recruited participants until the sample size met thematic saturation and exhaustive coverage of the twenty hospitals (37,38).

#### Data collection

We collected data in the respondent’s preferred language using audio-recordings and field notes. To ensure a culturally sensitive, women-centered environment, women’s IDIs by male researchers were conducted in the presence of a female community health worker, and FGDs in the presence of a local female midwife. They provided translations and contextual support when needed.

### Data processing and analysis

#### Data processing

We anonymised recordings and verified initial transcripts for accuracy. We did not translate the transcripts which were in French (majority) and English (minority) and did not return them to participants for comments or corrections.

#### Data analysis

Using NVivo® 14 software (Lumivero, LLC,) JTF and WCDK inductively coded the first four transcripts of each group of participants and built a harmonized codebook by merging their individual codebooks. They constantly memoed, refered to field notes, and triangulated by cross comparing. Saturation was reached with respect to the processes of IC and debriefing after coding the transcripts of 13 FGDs and 16 IDIs with post-CS women, but new meanings related to contextual socio-cultural features kept emerging, leading us to continue data collection and coding for all the remaining hospitals. Likewise, we coded all the transcripts of IDIs with CS providers (37,38). JTF edited the harmonised codebook that was reviewed by VF, MS, LTD and MM. Building on the final codebook, we generated themes through analysis of emerging patterns and identification of relationships.

## Results

### Sample characteristics

We conducted 20 FGDs with 128 pregnant women and 20 IDIs with post-CS women of whom 10 were interviewed together with their accompanying relatives (Fig. 1). In addition, we conducted 69 IDIs with CS providers. Table 1 summarises participants’ characteristics.

**Fig 1.**
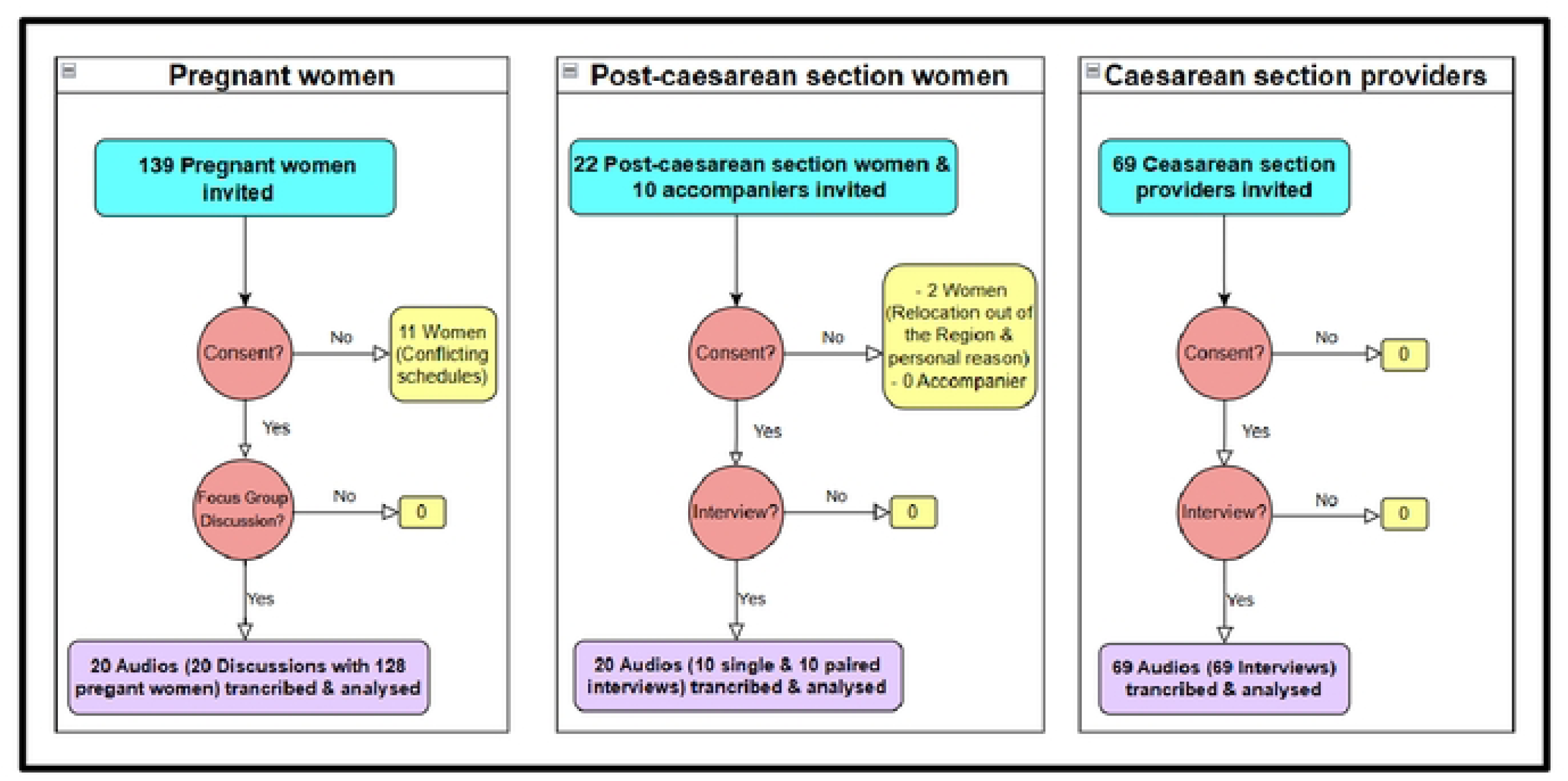
Flow diagram: inclusion of participants.

**Table 1.**
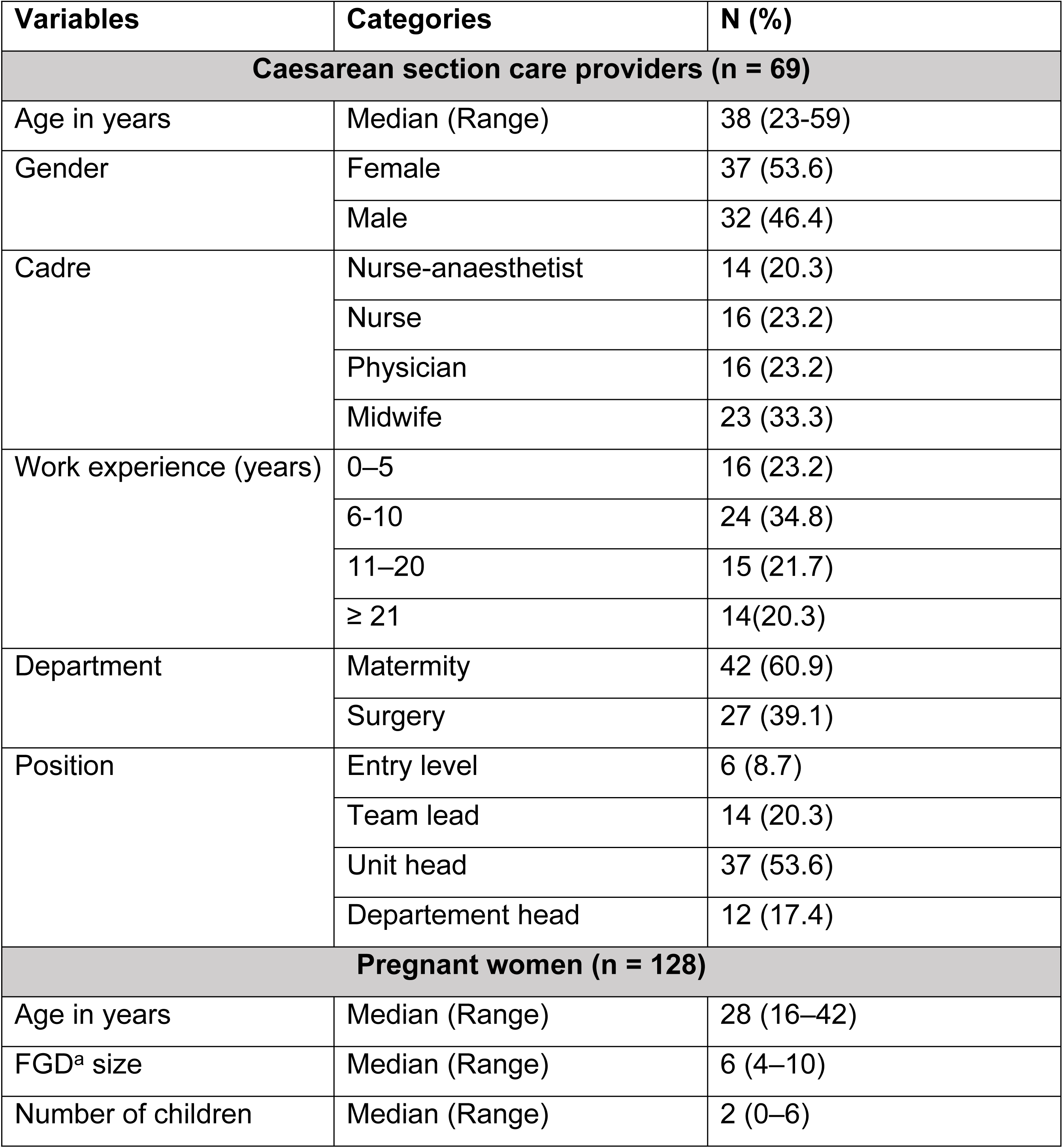
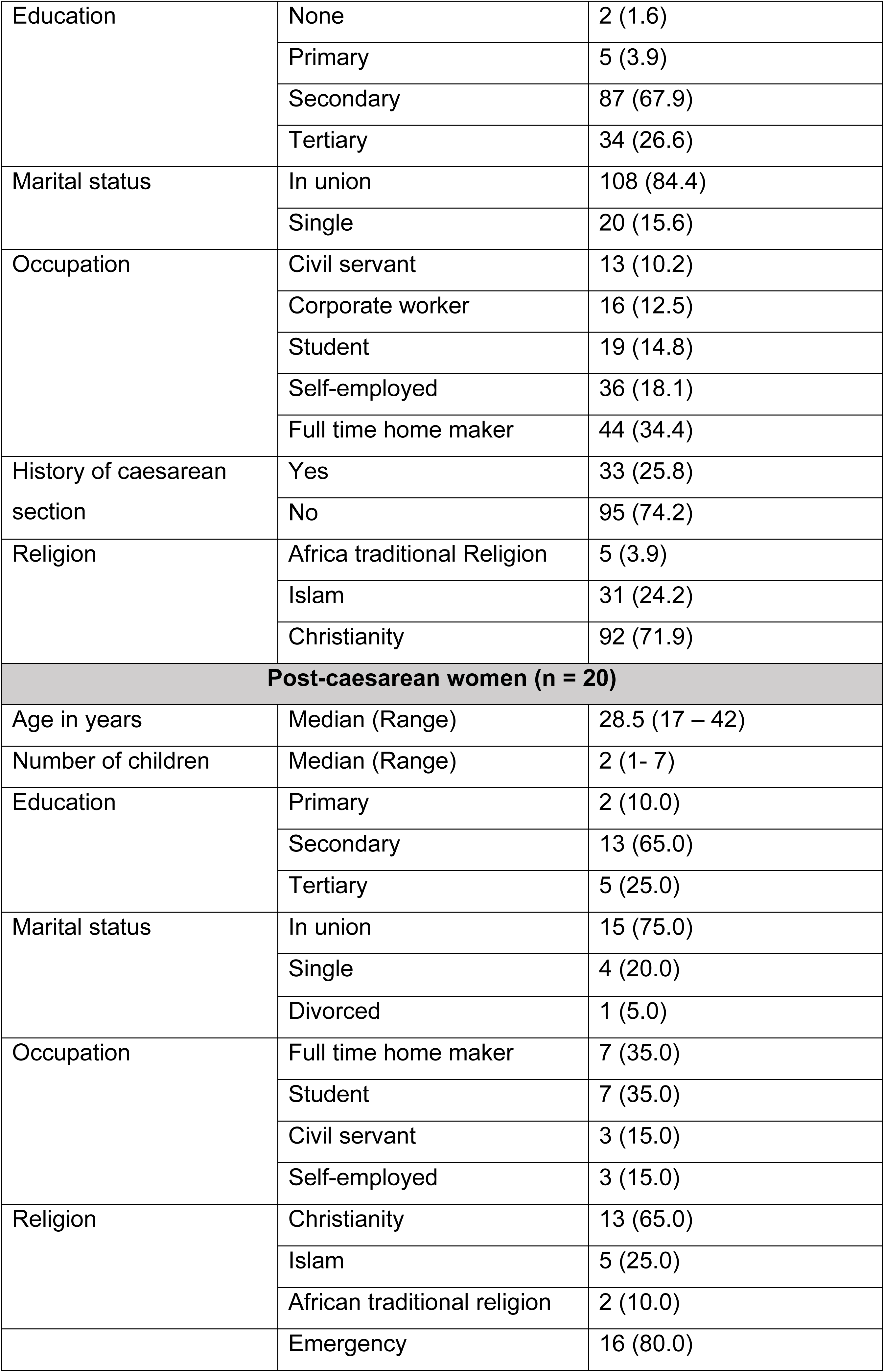
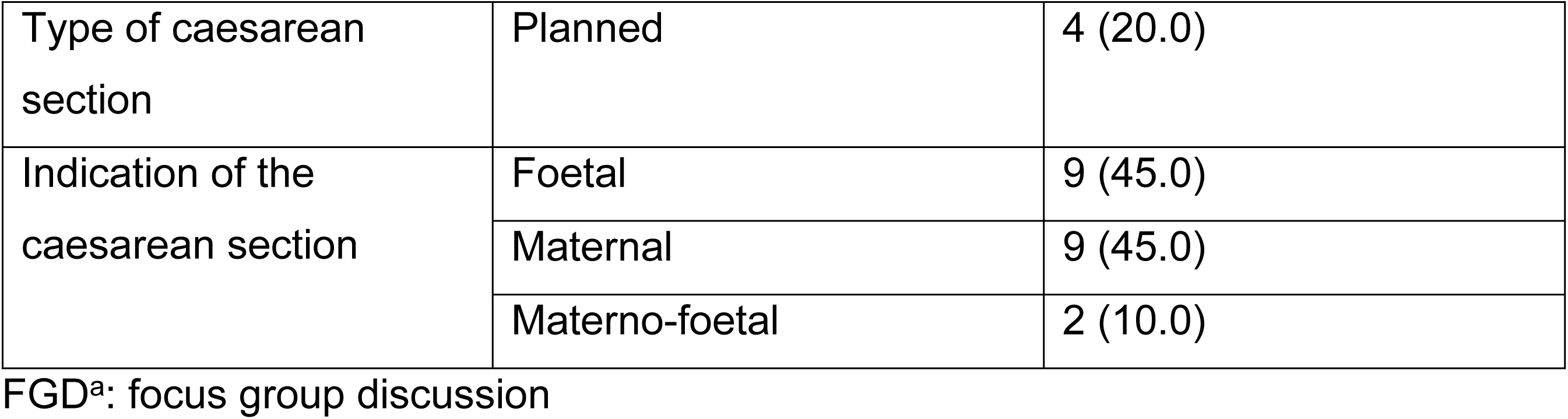
Characteristics of participants.

CS consent and debriefing processes in the Region are described elsewhere (39). Consent for elective CS is sought during antenatal consultations by the attending midwife from the woman who always secures her husband’s approval before notifying her final decision. In emergency CS, providers mainly engage with accompanying relatives who in turn refer to husbands, sometimes completely bypassing the woman herself. Debriefing after CS is not done in public hospitals and is content-limited in private faith-based hosptials.

### Themes

From the 109 transcripts, we inductively generated six themes. Five themes reflected barriers to women’s decision-making autonomy:

- primacy of the emergency context in CS decisions;
- women’s embeddedness in a patriarchal and collectivist society;
- medical paternalism and women’s deferential behavior;
- funding mechanism and logistical provision of CS care;
- weak regulatory framework for clinical practice.

In contrast, the sixth theme was a bold, emancipatory counter-current, challenging the above barriers and promoting women’s autonomy.

### The primacy of the emergency context for CS

CS providers said the high proportion of emergency CSs in the region shaped their approach to consent. Faced with women suffering life-threatening complications and/or labour pains, providers often assumed consent was *ipso facto* waived, implicit, or tacit. They consequently prioritised approval by accompanying relatives rather than women themselves. This routine bypassing of women eroded their autonomy and culminated in situations in which women were not told they were being taken to the operating room.

*“We think that because the woman at that moment will not have good decision-making ability … due to pressure. So, the family [members] are the ones to take the decision, the husband especially. At times there are some women they are in a state they cannot even speak. So, we don’t even bother to ask them”. A midwife*

*“When there is already an urgent indication, we inform the family. Most of the time, we don’t even inform the lady herself. It’s the family that comes now, we ask for their decision. If they don’t agree, we don’t go [i.e proceed].” A midwife,*

CS providers presumed women’s accompanying relatives were their “trusted representatives”. Even with that assumption, they didn’t meet the expectations of accompanying relatives who decried the short decision-making timeframe allocated to them or the lack thereof. They complained about being pressured to approve emergency CSs with no room for questions.

*“She [health worker] told me that I choose; that I sign. Because where the baby is, she [the woman in labour] can’t continue in the delivery room. We must go to the operating room. If I accept, they operate. If I refuse, they send her to another hospital. That’s when I signed.” An accompanying relative*

Similarly, post-CS women acknowledged that anxiety, stress and fear in emergency settings reduced their capacities for rational considerations. For some, it had been appropriate for CS providers to set the details of the emergency procedure with their relatives. Nevertheless, they emphasized that obtaining their own approval was crucial (albeit expedited or verbal given the emergency) prior to engaging with others. Moreover, they expected CS providers to retrospectively deliver the consent components skipped because of the emergency, during postoperative debriefing. The nearly systematic absence of debriefing deprived women of postoperative information which autonomous patients are entitled to after surgeries.

### Embeddedness in a patriarchal and collectivist society

Consent and debriefing interactions were not immune to social norms. In line with the prevailling patriachal and collectivist ethos, after the unsystematic request of women’s explicit, tacit or implicit consent, CS providers always sought an additional approval from a third party. This additional approval served as an endorsement by the woman’s support network. Most often, that power was vested in husbands (or fathers for unmarried women), who could veto both planned and emergency CSs. Such situations often forced providers into long and sensitive face-to-face or phone negotiations.

*“For the operation, I was surprised. In the operating room, they told me they had already agreed it with my husband. I don’t know why they didn’t tell me beforehand. … I don’t know what their intention was in keeping it from me. But when I arrived, I found myself ambushed like that; they told me: ‘Your husband has agreed.’ But I asked, “why am I being operated?” It was in the operating room that I was told that they couldn’t clearly hear the baby’s heartbeat and that they had to operate. I didn’t even have a choice.” A post-CS woman*

*“The purpose of consent is that we should not take a human being and put them on the operating table without their family’s approval… I ask their family, their husband… and they sign and give their phone numbers.” An Anaesthetist Nurse*

As we elaborate in the next paragraph, in emergency contexts CS providers often bypassed women and sought consent from husbands or fathers, either directly or through accompanying relatives. In some cases, husbands themselves sought approval from their wives’ fathers.

*“The husband will say: ‘No, wait, let me call first.’ He calls the woman’s family. You can feel that he doesn’t want to take full responsibility. He says, ‘It’s true that I’m with her, but her family must know that you want to perform a CS. I can’t carry this alone.’ So, this is always common. There’s always this fear of signing.” A General practitioner*

Women’s handling of their right to self-determination regarding CS reflected a substantial dependence upon their support networks and deference to men (husbands or fathers), either directly or indirectly, often through the intermediary of “*matrons*” (senior married mothers). In general, before consenting to a planned CS, women deliberated with their networks and sought approval from their husbands and/or elder women in their own families. When women did not get approval from their support network, antenatal consultation providers became involved by inviting husbands and matrons for in-person negotiations.

*“It’s true that she often agrees. And when she agrees knowing that the family will not support the decision, she already tells us that when we will tell her in-laws, they won’t agree. … When she accepts, she suggests that we speak to her in-laws ourselves. And sometimes, she even tells us exactly what to say.” A Midwife.*

*So, in situations where we say, humm, here’s what’s wrong, we’ll need to operate,’ we explain it, she tells you, ‘No, you need to tell this to my husband.’ Or even if the husband isn’t around, here’s my mother-in-law or here’s my mother. There’s always a third party who is there, it’s not really the woman’s decision.” A Midwife*

Older married pregnant women were stronger proponents of a patriarchal approach to consent and debriefing. They advocated that, as heads of households, husbands could righteously oppose an elective CS accepted by their wives, forcing CS providers to choose between implementing the husband’s decision or convincing him of the worthiness of the CS. A handful of post-CS young women shared that view, citing ancestral marital norms as essential to stable unions able to withstand the social and financial “turmoil” caused by CSs. Nevertheless, citing an Islamic law interpretation, one male respondent specified that a husband’s veto only applied to a medically unnecessary CS requested by his wife.

*“Once a woman has already received a dowry, she must not accept what her husband has refused—even if it’s a caesarean section, regardless of the consequences. She must rather convince him.” A pregnant woman*

*“According to the Muslim religion, a husband has full veto power only over elective caesarean sections on a woman’s request, and not over life-saving caesarean section.” A husband of a post-CS woman*

CS providers had so profoundly incorporated patriarchal norms in their approach to consent that they extended the omission of women’s consent to CS-associated procedures. Bilateral tubal ligation (an irreversible contraceptive method) is one of those procedures. According to Cameroon’s clinical practice guidelines, obtaining the woman’s consent well in advance is an absolute requirement. Nevertheless, a physician reportedly performed a bilateral tubal ligation during a fourth planned CS without the woman’s knowledge. He agreed with the husband not to inform the woman to spare her anxiety.

*Husband: “The doctor even called me; he told me that, no, the fourth caesarean was too risky and that it is necessary to tie the tubes.” Researcher: “Madam, did he tell you the same thing? Did they tell you at the same time?” Husband: “No. When he said that my wife wasn’t there, because he knew he couldn’t tell her that alone—it would have caused her stress.” (Everyone laughs) — A husband and his post-CS wife*

### Medical paternalism and women’s deferential behavior

CS providers exhibited paternalism both in consent transactions and in the near-total absence of debriefing. This was fuelled by two provider-related drivers and a women-related one. Firstly, providers expressed a sense of “saviourism” toward distressed or agonising women-baby dyads referred with life-threatening pregnancy complications – often following failed care by insufficiently skilled staff at lower-level health facilities or by non-conventional community health providers (traditional or faith-based healers).

*“Well, those who come from the community rather praise us because they arrive already dying. They say to themselves that they’re just going like that, to be sent to the mortuary. When we manage to save them, they realize that we are quite effective.” A midwife*

*“We are in rural zones. So, where we are, people remain archaic. They are closed-minded, taboos and all. They belief that staying in the neighbourhoods drinking those potions it [foetus] will come out by itself. They believe that magic exists. They believe that by going there for [childbirth] labour if they operate me, I will not get up [from the operating table]. It is because they are closed on themselves and don’t believe in modern medicine. They [women], believe much more in traditional medicine.” A midwife*

Secondly, antenatal care providers appeared to have limited proficiency in women-centred discussion of the possibility of CS. Both junior and senior CS providers revealed that such competencies were neither thoroughly taught during pre-service training nor integrated into routine in-service training.

*“May this [research] project help us really get training in counselling women during antenatal care, in the delivery room, and for those who are going for caesarean sections, that would be really good. That’s missing in the hospital. All of us here, this is what we need We don’t think about counselling. … We talk about it, but we don’t do it. And we fail because we don’t do it.” A midwife*

Thirdly, women’s deferential attitudes regarding CS providers reinforced medical paternalism. Most women exhibited little individual agency in interactions with CS providers, as mentioned earlier routinely deferring to husbands or in-laws for consent decisions. For these same reasons, post-CS debriefing with women was not routine. Providers often did not view women as entitled to explanations and women seldom felt able to request them. When they did, responses were vague or ignored, and at times key details were shared with relatives without women’s consent.

*“In that hospital, if you don’t ask questions they [CS providers] will explain you nothing about your cesarean section. Apart from injections and tablets, they just make sure you pay your bill before you leave” A post-CS woman*

*“I did not have any communication after the caesarean…. So, the few pieces of information I got were the ones I forced myself to get. There is no communication. If*

### you can’t go towards information, the information will never come to you.” A post-CS woman

*“Do you ask a woman before inviting a third party in the debriefing? Generally, we don’t ask the woman. In principle, if she has already told us, ‘Here is my husband,’ we explain to him what was done in the operating room and the difficulties we encountered.”*

### An Obstetrician Gynaecologist

Most women insisted that the best way to guarantee a conducive domestic environment for post-CS recovery (e.g. wound care, restrictions on lifting heavy objects, sexual intercourse, farm work) was for CS providers to talk to husbands on the day of discharge.

Thus, women’s deference towards CS providers and husbands significantly undermined their autonomy.

*“When the doctor says it, no one can’t say that you made it up or that you are avoiding your marital responsibilities [household chores, farm work, sex….]. It prevents a lot of conflict at home.” A pregnant woman*

### Funding mechanism and logistical provision of CS care

The CS-funding model alongside women’s (lack of) financial autonomy, emerged as a major driver for autonomy-undermining practices. Compulsory out-of-pocket payments before planned CSs, and early post-operative payments for emergency CSs, made the ability to pay a *sine qua non* requirement for consent. Where consent forms were used, the signature had a dual meaning: consent to CS and commitment to pay.

*“Now, the difficulty is often with emergency caesareans. Because in emergency caesareans… what we often notice is that the person who pays has the final say. This means that to get authorization, sometimes you have to call up to four people.” A midwife*

Across all respondent categories, lack of insurance, universal health coverage and women’s inability to pay for CS from their own income and their near-total reliance on their husband or parents was a key determinant of their dependence on them for consent.

*“He [husband] is honoured (laughs) … because he is the father of the child. …. (laughs) If there are bills, we give them to him. Even if your mother has the money, she can’t pay without agreement of your husband.” A pregnant woman*

*“Often, these are young girls under the authority of their parents. So, they wait mostly for the parents’ approval. For women under the financial authority of their husbands when she wants to accept, the caesarean section, the husband’s authorization is needed because he is the one who pays at the end.” An Obstetrician Gynaecologist*

Consequently, husbands are decision-makers at a critical juncture for their wives’ health and wellbeing. Women made it clear that husbands’ decisions did not always align with the medical need for CS. Furthermore, it is customary for husbands to be absent or in secondary roles during childbirth, leaving their mothers to take their daughters-in-law to the hospital and oversee the process. In general, women blamed mothers-in-law for their reluctance toward necessary CSs because of financial reasons.

*“If she falls onto the mother-in-law, she will say No, my son doesn’t have that kind of money. And please, that woman is unlucky. Among all the women out there, he had to find an unlucky woman who will give birth through caesarean. Is it even certain that it’s a caesarean the doctors are requesting over there? She’s the one who requests it. She wants to empty my son’s pockets!” A pregnant woman*

Logistical aspects of CS also reduced women’s autonomy, albeit to a lesser extent. Structural staff shortages force patients to have a carer throughout their hospital stay. This crucial role is performed by close relatives and encapsulates purchasing emergency supplies often from outside the hospital and caring for the newborn. It is therefore relevant for CS providers to gain the carer’s full cooperation by involving them as co-consenters. In that capacity, they were sometimes very effective allies in persuading women to accept the prescribed emergency CS, although most of the time they were the actual consenters. In situations where accompaniers opposed the CS, providers complained of spending precious time convincing them before conducting the procedure, regardless of the woman’s opinion.

### Weak regulatory framework for clinical practice

In over two-thirds of hospitals, CS providers reported the absence of guidelines to standardise consent and debriefing. Consequently, they relied either on informal consensus within units or on individual discretion. The lack of standardised forms in most hospitals, combined with providers’ limited skillsets, caused alignment of consent practices on prevailing professional routines (medical paternalism) and community norms (third party’s consent). Where a standardised consent form was available, its utilization was suboptimal or spurious. Suboptimal practices included obtaining signature from women and their relatives with little or no time for reading it or requesting signature after the CS merely to fulfil procedural requirements.

*When there is an emergency, it is the accompanier who signs because the woman in labour is in an urgent situation. It is after she leaves the operating room that she is asked to complete the formalities because it is also her duty to sign.” A midwife*

*“Researcher: They made you to sign. Did you read what you signed? Accompanier: They only said I sign; I put my identity card number, and we go to operating room.” An accompanying relative*.

Sometimes, the process was outright spurious: CS providers listed as signatories had not seen the form. Besides, no hospital gave women a copy of the signed consent form.

*“It’s true that consent forms are there, but I don’t even know when they are filled out. Afterwards, I just see my name, stating that “I talked with the doctor”, as they say, though I never actually spoke with the patient.” A general practitioner*

### Bold emancipatory counter-current

We identified a bold emancipatory trend towards women-centred consent and debriefing, spearheaded by younger women and backed up by a minority of CS providers (mostly seniors and executives). They openly condemned and challenged prevailing socio-marital norms and care delivery routines. The overarching principle of that counter current was that no third party should patriarchally or financially stand in the way of women’s and child’s health benefits when it comes to CS, regardless of relationships or vested interests at stake. Those advocates substantiated their positions with three arguments. Firstly, women insisted that providers must always seek and uphold women’s consent irrespective of the opinion of their support networks; they insisted that husbands’ and accompanying relatives’ roles should be limited to supporting women’s choices

*“Well, in my opinion, I would first talk with my doctor. If it concerns me, I don’t see why my mother or my husband should be informed before me.” A pregnant woman*

A few CS providers revealed that they followed this principle, conducting emergency CSs against husbands’ refusal to pay after a woman’s commitment to pay soon after surgery.

*“I have already taken it upon myself to admit a woman to the operating room for an emergency caesarean despite her husband’s refusal, after the patient committed to paying herself after leaving the operating room.” A nurse anaesthetist*

Some women extended this radical stance on women’s bodily autonomy to birth spacing, strongly advocating for husbands to sign pledges for a minimal interpregnancy interval after CS.

*“What I mean is, if he doesn’t understand me regarding the birth spacing that has been recommended and I end up getting pregnant too soon, if it costs me my life, for example—after me, he’ll remarry once again. Who will lose? It’s me. That’s why I prefer that it should be written down, and I sign, and he signs too.” A pregnant woman*

Secondly, acknowledging that the out-of-pocket funding model undermines women’s autonomy, both women and CS providers urged the government to accelerate the roll out of universal health coverage to include every CS, thus reducing their dependence on husbands.

*“What I’m asking the president is lower cost of caesarean sections for all women in the country. Because it’s not right: a man gets a woman pregnant, and then we’re talking about surgery, and he must pay 400’000 [xaf] francs [714 USD]. That’s impossible.” A pregnant woman*

Thirdly, these women firmly stated they would undergo a CS against their husbands’ will, even at the cost of their marriage and hold CS providers accountable for disregarding women’s rights to self-determination. They also wished more women would do the same in future.

*“I must accept the emergency caesarean even if the husband refuses and divorce after rather than dying of having labour complications. He will take many other wives you have only one life.” A post-CS woman*.

## Discussion

Women’s autonomy in consent and debriefing for CS lies at the intersection of two contradictory forces: the imperative to uphold “women-centredness” as a fundamental principle of high quality of care and the persistent influence of gender-based discrimination in everyday practices within hospitals. We found that autonomy was gravely undermined by CS providers, women’s support networks and women themselves. However, despite these autonomy-inhibiting mechanisms, there was an emancipatory trend, which identified financially prohibitive cost of CS as key drivers of disempowerment of women in this process.

Women’s lack of agency regarding informed consent and debriefing was shaped by their limited financial capacity by internalized deference mechanisms towards their husbands. In Cameroon, only one in two married women participated in decision-making regarding their healthcare in 2013-18 (34). Our findings are novel in showing the depth of these deferential attitudes, which persisted despite the high health stakes of CSs.

Moral philosophers who posited authenticity as a prerequisite for relational autonomy have explained such a lack of self-esteem and self-trust by the working of deeply internalised domination schemes (1). They argued that autonomy is inherently social-relational, as individuals always deploy self-determination in relation to fellow.

Authenticity or trueness to oneself will foster autonomy only if the agent possesses enough self-respect and self-esteem which is then matched by their voluntary undertakings. Individuals (such as subservient minorities, slaves, or deferential wives) who failed to develop enough self-esteem because their characters were shaped in social, cultural and educational systems of institutionalised oppressions or dominations can hardly exhibit auto-determination in a way that trumps their internalised conformity patterns.

Similar patterns were found in Nigeria, where Ugwu and De Kok linked CS refusal to patriarchal domination and financial dependence, while Enabudoso and Igbarumah found that half of women who underwent CS preferred their husbands to sign the consent form (40,41). Our findings confirm that financial dependence reinforces husbands’ authority rooted in marital norms (36,42). In contrast, women in our study demanded for husbands’ commitment over post-CS contraception, which is free of charge, sometimes even suggesting that the husbands provide written pledge in hospitals. This could indicate a relatively high level of family planning awareness, but also the emancipatory action of financial autonomy (43,44).

While in both individualist and collectivist societies institutionalised oppression systems are exerted through political and economic structures clearly external to individuals, domination patterns easily permeate collectivist societies masquerading as self-enforced conformity to socialized power and norms (45,46). The semi-bantou and bantou ethnic groups across sub-Saharan Africa share a collectivist trait reflected in the ubuntu philosophical maxim “I am because of who we will all are” (Mugumbate and Andrew, 2013). In West Cameroon, collective norms include deep respect for the elders and hierarchy, deferential greetings, respectful body language, elder-led and men-led decision-making and men’s leadership and financing of households (36,48). Husbands’ and CS providers’ paternalistic behaviours were possibly unopposed because women’s thoughts and will align with deference and respect towards hierarchies. Husbands enacted their power of heads of household while in the absence of compelling practice guidelines CS providers embodied a position of power in their maternity units. Indeed, both public and faith-based private hospitals project the power of their parent institutions, namely the government and Christian churches, which are highly respected in Cameroon (49).

Women’s autonomy was also restricted by structural and process factors cross-cutting several WHO’s health system building blocks (19,50,51)Severe staff shortages left CS providers overworked, prioritising clinical interventions over users’ experience-related services including consent and post-CS debriefing (25,26). The limited literature on burnout among Cameroonian health staff reveals a substantial prevalence (52,53). Structural factors also include providers’ inadequate skillset emphasized by CS providers who complained that their pre- and in-service education on informed consent, debriefing and psycho-emotional support was insufficient. In Ghana, midwives deplored similar limitations to women-centred care, namely staff shortage and limited in-service training (54).

Process factors were the absence of standard operating procedures (including lack of standardised forms) and the burden of out-of-pocket expenditures. Such payments are a well-known barrier to care. According to a recent review, at least thirteen sub-Saharan Africa countries have reformed financing mechanisms including reducing reliance on out-of-pocket payments and expanding health insurance (55). While these reforms aimed to improve access and reduce inequalities, free CS policies can also change providers’ behaviour in unintended ways. By removing the need for immediate financial commitment and without additional guidelines for consent, such policies may contribute to more minimal interactions between providers and women as observed in Benin (56). Anticipating and mitigating providers’ resistance or adaptive behaviour that may perpetuate autonomy-undermining practices will be essential when developing new policies to reduce the financial burden of CS on women and their families (56–58).

While trusted representatives can provide consent on behalf of women lacking decision-making capacity (mentally, physically or legally), the systematic presence of another person when adult competent women are making decisions about their CS questions the socio-medical handling of women’s autonomy. The issue is not whether CS providers should talk to accompanying relatives but the shifted locus of decision-making and frequent departure from women’s best interests. Beyond the universally agreed principle of uncoerced and informed women’s decision-making, the pluralistic praxis of medical ethics results in contextual derivations through socio-cultural contamination (7,59). In liberal individualist societies, it is easier to get women deciding on their own than in conservative and traditional societies because decision logics generally diverge from self-centred individual desires. Moreover, in some Islamic settings, physicians abide by religious obligations that restrict patient’s autonomy. For instance, the Islamic code of medical ethics refutes autonomy as grounds to contravene Islamic precepts in medical practice, emphasizing instead a balance between individual rights, family wishes and societal requirements (2). The dual loyalty of health staff to their individual patients and to the society could explain why CS providers systematically sought consent from husbands in the collectivist and partially Muslim West Region of Cameroon. The limited literature on consent for CS in sub-Saharan Africa shows a similar pattern in Nigeria (41,60). Nevertheless, this should not justify providers playing a role in undermining women’s autonomy. The World Medical Association recommends that when the interests of society conflict with those of the patient, physicians should stand and act for the patient (Ball and Corbis, 2015).

### Push for agency

It is meaningful that the movement toward women’s agency was advocated by both younger women and seasoned senior professionals with managerial roles. This tandem has major implications. First, on the community side, young women making up most of the female population in Cameroon would provide the critical proportion of early adopters of any policy strategy aiming at increasing women’s self-determination in hospitals (61,62). Second, on health systems side, the strict hierarchy in hospitals demands a high level of power which only senior or highly ranking staff have, to contravene the organizational routine of driving away or referring women to other hospitals when husbands or accompaniers opposed a medically indicated CS (Howard-Grenville and Rerup, 2016). These routine practices could be labelled as institutional deception towards women given that their healthcare is disregarded to satisfy third parties (64). Our findings suggest that provider’s leadership can change the paternalistic, patriarchy-leaning and non-standardized consent and debriefing processes. Indeed, peer-initiated changes face less resistance and are more likely to be adopted by front-line providers (65).

### Implications for policies and future research directions

Policymakers should develop and disseminate standard operating procedures for consent that foster women’s agency. This includes ensuring that CS providers explore each woman’s perspective, amplify her voice, and remain guided by her wishes. Debriefing after CS should also centre on the woman needs and provide her with enough support to secure the adherence of her social network to her preferences. Community engagement should factor in the deeply rooted influence of marital norms and women’s limited financial capabilities on CS-related decision-making. Finally, social protection mechanisms should cover every CS to alleviate financial constraints that perpetuates women’s dependence on their husbands. In the era of universal health coverage, implementation research could examine how removing users’ fees for CS might enhance women’s autonomy in giving consent.

### Strengths and limitations

Interviews and analyses by an obstetrician and a social scientist captured both biomedical and socio-cultural aspects. We collected data in most socio-cultural contexts of the Region. We explored both the supply and users’ sides. In every interview, a female member from the local community participated, helping foster trust and authenticity through cultural acceptability.

On the other hand, most of the quotations are translations from French that may have failed to convey original meanings despite careful translation. Interviewing providers inside their workplaces could have led to self-censorship which was mitigated by triangulation and familiarity given the clinical background of the lead interviewer. The presence of local female health personnel during IDIs and FGDs with women might have triggered a desirability bias silencing harshest criticism. To avoid this, we briefed our female collaborators and encouraged participants to elaborate on every apparent or latent criticism. We did not directly observe real-life consent or debriefing, missing potentially informative unconscious behaviours. Finally, the presence of accompanying relatives in half of women’s IDIs may have tempered their criticisms against patriarchy.

## Conclusions

This study revealed that women’s autonomy regarding consent for CS and debriefing across the West Region of Cameroon is gravely undermined by women themselves, women’s support networks, CS providers, and health systems shortcomings. Nevertheless, a bold emancipatory current, driven by younger women and senior CS providers, challenges the prevailing status quo and pushes towards increased women’s autonomy. Decisive actions are needed in hospitals and communities to foster women’s autonomous decision-making in informed consent and debriefing for CS.

## Data Availability

Data Supporting this article, namely the transcripts of in-depth interviews and focus group discussions are available upon reasonable request to the corresponding and senior authors.

## Acknowledgements

We are grateful to the regional sexual and reproductive focal person and to the Regional Delegate for Public Health of the West Region of Cameroon for their support and advice. We would also like to thank the participants who generously lent their time to this research.

## Notes

### Competing Interest Statement

This manuscript is original work and has not been published or submitted elsewhere. All authors have approved this submission and have no conflicts of interest to declare.

### Funding Statement

This research was supported by the Japanese Government through its world innovative and smart education grant, and by the London school of Hygiene and Tropical Medicine through its research degree travel grant. The funders had no role in study design, data collection and analysis, decision to publish, or preparation of the manuscript.

### Author Declarations

LSHTM Research governance and integrity Office Regional Ethics Committee for Human Health research of the West Region of Cameroon

